# Understanding problems and solutions related to accessing cervical screening for people with a physical disability, condition, impairment or difference

**DOI:** 10.64898/2026.05.27.26354052

**Authors:** Emma Kemp, Julius Sim, Katie Wright-Bevans, Samantha Renke, Carolyn A. Chew-Graham, Andrew Finney, Charlotte Harper, Laura A.V Marlow, Susan M. Sherman

**Author notes:** Corresponding author: Susan M Sherman, Address: School of Psychology, ICOSS Building, 219, Portobello, Sheffield, S1 4DP.

## Abstract

**Background:** Physically Disabled women are less likely to access cervical screening than non-disabled women, yet little research has been conducted to understand the problems that Disabled women face or potential solutions.

**Methods:** A cross-sectional online survey was conducted with 1493 UK-based participants who identified as having a physical disability, impairment, condition, or difference that makes cervical screening difficult or impossible. Participants were presented with statements about cervical screening problems and potential solutions and asked to indicate agreement using a 5-point scale. They also provided open-ended comments. Data were analysed using descriptive statistics, multinomial logistic regression and thematic analysis.

**Results:** More than half of participants reported delaying/missing (46.8%) or never attending (8.8%) screening, with most of those (71.0% and 81.4% respectively) indicating that the main reason was disability-related factors. The highest levels of agreement for problems were for concerns about pain, embarrassment, and fear of what the test might find and for potential solutions were for having a doctor or nurse who is willing to try different solutions, discusses specific needs, and understands physical disability. Never-attendance (OR = 0.022, 95% CI 0.014, 0.035) and delaying or missing appointments, (OR = 0.057, 95% CI 0.043, 0.076) negatively predicted future screening attendance. Six themes were identified from open-ended comments, supporting and extending the quantitative findings.

**Conclusion:** Disabled women face the same problems related to cervical screening as non-disabled women and additionally face disability-specific problems. Cervical sample taker training should incorporate ways to support physically Disabled women to have equitable access to screening.

## Introduction

Cervical cancer was responsible for 348,189 deaths globally in 2022 and is the fourth most common cancer in women worldwide (Bray et al, 2024). The World Health Organization has outlined a global strategy to eliminate cervical cancer (defined as 4 or fewer cases per 100,000-woman years), which includes ensuring that 70% of women receive cervical screening by 35 years of age and again by 45 years of age (WHO, 2020). A considerable amount of research has been conducted to understand the factors that influence cervical screening uptake, with a view to increasing uptake and reducing inequalities (e.g. Shpendi et al., 2025; Wearn & Shepherd, 2024). However, very little research has been conducted to understand the factors that influence cervical screening uptake for physically Disabled^1^ women and people with a cervix (henceforth ‘women’).

Physically Disabled people face numerous inequalities in healthcare, such as inaccessible environments, lack of height-adjustable examination tables, and healthcare professionals’ limited understanding of how to accommodate physical differences (e.g. Andresen et al., 2013; Smeltzer, 2007; Iezzoni et al., 2016; Vinson, 2025). In a recent scoping review, Harper et al (2026) reported that physically Disabled women do not engage in cervical screening as often as non-disabled women. Despite there being an estimated 700 million Disabled women and girls globally (UN Women, 2024), a recent systematic review identified just nine papers exploring factors that influence cervical screening uptake for physically Disabled women (Chan et al, 2022), none of which were conducted in the UK. This reveals a considerable gap in health equity research, yet equitable cervical cancer elimination is impossible unless elimination is achieved for everyone.

In the UK, there are currently over 8 million Disabled women, with 41% of Disabled working-age adults reporting mobility issues (Department for Work and Pensions, 2025). Cervical screening is available to women aged 25–64, who are invited every five years as part of the NHS screening programme (NHS UK, 2025), saving an estimated 5000 lives annually (Peto et al, 2004). However, inequalities persist. Jo’s Cervical Cancer Trust (2019) investigated barriers to cervical screening for 335 physically Disabled women in the UK, reporting that 88% found it hard to access or attend cervical screening, 63% were unable to attend due to their disability and a lack of access and equipment at their GP surgery, and 49% did not attend due to previous bad experience or concerns around staff attitudes. Chan et al’s (2022) review revealed that challenges in accessing screening providers and undergoing cervical screening, uncomfortable experiences during the screening procedure, and a lack of knowledge of cervical screening and how it can be accessed were major impediments to accessing and attending cervical screening.

The aim of this study was to investigate problems faced by physically Disabled women in the UK regarding cervical screening uptake and identify potential solutions to make cervical screening less challenging. This was explored through two research questions:

1: What problems and solutions are most frequently endorsed?

2: What factors affect the likelihood of future cervical screening attendance?

## Method

### Design

We conducted a cross-sectional survey that explored current screening attendance and problems, solutions and preferences regarding screening. This survey also explored attitudes and preferences relating to human papillomavirus (HPV) self-sampling (reported separately; Kemp et al, 2026). We conducted the survey using two recruitment approaches: i) an online panel (<u>Prolific.com</u>) and ii) charities advertising and existing networks. The data were collected between July 2024 and May 2025. The survey was delivered on the online survey platform Qualtrics.

### Patient and Public Involvement

One of the co-authors, SR, was the patient and public involvement (PPI) lead for this project. She has osteogenesis imperfecta and is a full-time wheelchair user. We additionally recruited 14 physically Disabled women (ages 26–63) to form our stakeholder advisory group. SS and EK met regularly with SR, meeting with the stakeholder advisory group three times to develop the materials. We presented the advisory group with a first draft of the survey, based on previous research. They generated additional items based on their own experience and then rank ordered them by importance to form the final survey items. They also helped us to develop the recruitment method, whereby potential participants were able to self-identify their physical status, and assisted with publicizing the study. The advisory group was invited to comment on a draft of this manuscript and their comments have been incorporated and acknowledged.

### Participants

#### Eligibility criteria

Participants had to identify as having a physical disability, impairment, condition or difference that makes cervical screening difficult or impossible, be aged 25–64 (screening age), be living in the UK, and to have been invited to cervical screening (even if they did not attend). Participants were not eligible to take part if they had received a total hysterectomy. Those who took part via the online panel had to first complete a brief screening survey to assess eligibility before being directed to the main survey. The main survey assessed the eligibility criteria, and this served as the screening questions for those individuals recruited through charities.

#### Online panel recruitment

The first wave of participants (*n*=1000) was recruited in July 2024, with a second wave of recruitment (*n*=439) in March 2025 due to low uptake of recruitment using alternative methods. Participants were paid £0.10 for completing the eligibility survey and £3.00 for completing the main survey. For the main study, participants who failed more than one attention check question (out of four) were not reimbursed and their data were excluded from analysis.

#### Charity-based/PPI links recruitment

To ensure representation of a range of disabilities and conditions, we aimed to supplement data collected via the online panel by recruiting people through relevant charities (e.g., SCOPE, Spinal Injuries Association, Brittle Bone Society) and our PPI stakeholder members’ personal networks. Participants recruited through this method (N=54) completed the main survey on Qualtrics and were reimbursed with a £5.00 voucher. We were unable to collect more participants through this route due to repeated hacking of the survey, despite the many controls in place.

### Measures

The full survey is available at: https://osf.io/fbmd2. Participants were presented with an information sheet outlining the nature of the study and were required to provide online consent.

For the main survey, we included four items to capture how participants describe their physical disability, condition, impairment or difference. We asked: ‘Please can you tell us the nature of your physical disability, condition, impairment, or difference’ (optional open-ended question). Participants were then asked: ‘Does your physical disability, condition, impairment or difference affect you in any of the following areas?’. This question was based on the 10-item Impairment Harmonised Standard (Government Analysis Function, 2020), with additional items adapted from the International Classification of Functioning, Disability, and Health (World Health Organization, 2003) to include pain, sexual function and urinary function. Participants were then asked if they used assistive devices. If they answered ‘yes’, we provided them with a list of options from Melbourne School of Population and Global Health (2022).

Six items modified from Jo’s Cervical Cancer Trust (2019) survey relating to cervical screening status were provided to measure participants’ previous experience of, and future intentions to attend, cervical screening. We asked participants ‘Which of the following best describes your previous participation in cervical screening?’ with response options: ‘I have never attended’, ‘I have attended but sometimes delayed or missed my screening appointment (more than 6 months)’, ‘I have always attended when invited (within 6 months)’. If participants selected ‘I have never attended’ or ‘I have sometimes delayed or missed screening’, they were asked if this was due to their physical disability, condition, impairment or difference. Participants who indicated that they had previously attended screening were asked to select where they had the test. All participants were then asked to indicate their likelihood of attending screening in the future with a 5-point scale (very unlikely to very likely).

Participants were presented with problems derived from previous research (Waller et al., 2009, Jo’s Cervical Cancer Trust, 2019) and discussions with our PPI group, who identified additional problems based on their own experience of cervical screening. They were also presented with solutions to cervical screening attendance adapted from the Jo’s Cervical Cancer Trust (2019) survey and based on PPI discussion. To ensure accessibility, we provided participants with the choice of having the problems and solutions presented in a list or a grid format. We modified the wording from ‘smear test’ to ‘cervical screening’ based on PPI preferences. Participants were asked how much they agreed or disagreed with each problem statement and to what extent they agreed that each item would (or already does) make cervical screening easier for them to attend on a 5-point Likert scale (strongly disagree to strongly agree).

Participants were also provided with open-ended text boxes and asked if there are any other factors ‘that make it challenging or impossible for you to attend cervical screening?’ and ‘that do, or would, make it easier for you to attend cervical screening?’

Socio-demographic questions were asked to collect participants’ gender and ethnic background (wording from UK Census, 2021), location, religion, education (wording from YouGov, 2020), household income, relationship status, and language. Finally, we included two items to provide participants with the opportunity to write freely about their cervical screening experiences and provide comments on the survey.

A debrief directed participants to further support and advised participants to contact their GP if they have any questions.

### Data analysis

We adopted a mixed-methods approach that included statistical analysis of the quantitative questions and thematic analysis of the three open-ended questions relating to problems, solutions and cervical screening experience.

#### Quantitative data

Descriptive statistics were calculated for participant demographics, cervical screening practices, disability characteristics and problems and solutions related to screening.

To examine potential predictors of future screening, we used ordinal logistic regression. The outcome variable comprised responses to a question asking respondents how likely they were to go to cervical screening when next invited: ‘very unlikely’(1), ‘somewhat unlikely’(2), ‘neither likely nor unlikely’(3), ‘somewhat likely’(4), ‘very likely’(5). Variables representing demographic characteristics, previous screening attendance, aspects of disability, perceived problems with screening, and potential improvements to screening were specified *a priori* as predictor variables and included in the regression model in blocks (see Table 8). We reduced the 53 questionnaire items relating to the problems and solutions to 11 latent variables using principal components analysis (with varimax rotation and Kaiser normalization); see supplementary materials Tables A3 and A4 for the derivation of the components.

When comparing a category on a nominal predictor to the reference category, or when considering a one-unit increase on a continuous predictor, an odds ratio (OR) from the regression analysis indicates how much larger (OR greater than 1) or smaller (OR less than 1) the odds are of moving upwards by one level at any point (except the last) on the outcome variable. Here, an increase of one level on the outcome variable indicates greater likelihood of attending screening, so that, for example, an OR greater than 1 indicates an increased likelihood of future screening with a one-unit increase in the predictor variable. The magnitude of odds ratios can meaningfully be compared within, but not between, nominal and continuous predictors.

The explanatory power of the model was measured using the Nagelkerke pseudo-*R*^2^ statistic (range 0–1) (Nagelkerke, 1991), which denotes the degree to which values of the outcome variable can be explained by the predictors in the model. We measured the additional explanatory power of each block in the model by calculating the increase in the Nagelkerke *R*^2^ when the block was added to the model containing the other blocks (i.e. the difference in *R*^2^ value between the full and reduced models). The goodness-of-fit of the full model was examined through a deviance chi-square test (where a non-significant test indicates satisfactory goodness-of-fit) and the assumption of proportional odds was tested by calculating and examining separate ORs at each threshold. We calculated a *p* value for each predictor and set statistical significance at *p* ≤ .05 (two-tailed), with 95% confidence intervals derived for all ORs.

#### Qualitative data

A codebook thematic analysis approach (Braun & Clarke, 2023; Boyatzis, 1998) was used to analyse the open-ended responses related to problems, solutions and screening experience (see supplementary file for codebook). This method of thematic analysis offers a structured approach suited to applied research questions while permitting an interpretive analysis of the data (Braun & Clarke, 2022). After removing single word responses such as ‘no’ that did not contain any semantic meaning, we analysed 851 detailed responses relating to problems, 666 relating to solutions, and 580 related to screening experiences. Two coders (EK, KWB) took an inductive approach to coding and initially coded 15% of the data. After initial coding, inter-coder review took place, with discrepancies resolved. Once agreement had been reached, both coders began to develop the codebook. Coder one (EK) then continued to code the remaining data and complete the codebook. Initial themes were then created from the codebook and both coders met to develop these themes. These themes were also reviewed by a third (SS) and fourth (JS) coder. After review, six final themes and associated descriptions were developed.

## Results

1602 participants took part in the survey. Of these, 109 respondents were excluded due to incomplete survey responses (*n*=88), more than one failed attention check (*n*=8), or because of ineligibility (*n*=13). After exclusions there were 1493 participants included in the analyses. Participant characteristics are shown in Table 1. The dataset is available at: https://osf.io/ufx8r/files/tm2us.

**Table 1.** Participant characteristics.

|  |  |  |
| --- | --- | --- |
| Age; mean (SD) |  | 41.71 (10.96) |
| Gender identity; $n$ (%) | Male | 2 (0.1) |
|  | Female | 1464 (98.1) |
|  | Non-binary | 17 (1.1) |
|  | Other | 9 (0.6) |
|  | Prefer not to say | 1 (0.1) |
| Ethnicity; $n$ (%) | English, Welsh, Scottish, Northern Irish, British | 1333 (89.3) |
|  | Irish | 5 (0.3) |
|  | Gypsy or Irish traveller | 1 (0.1) |
|  | Any other white background | 26 (1.7) |
|  | White and Black Caribbean | 13 (0.9) |
|  | White and Black African | 8 (0.5) |
|  | White and Asian | 13 (0.9) |
|  | Any other mixed or multiple background | 11 (0.7) |
|  | Indian | 14 (0.9) |
|  | Pakistani | 7 (0.5) |
|  | Bangladeshi | 6 (0.4) |
|  | Chinese | 12 (0.8) |
|  | Any other Asian background | 1 (0.1) |
|  | Caribbean | 15 (1.0) |
|  | African | 10 (0.7) |
|  | Any other Black, Black British or Caribbean background | 4 (0.3) |
|  | Arab | 1 (0.1) |
|  | Other | 8 (0.5) |
|  | Prefer not to say | 2 (0.1) |
|  | Missing | 3 (0.2) |
| Main language; *n* (%) | English | 1487 (99.6) |
|  | Other | 6 (0.4) |
| Highest qualification; *n* (%) | Degree equivalent or higher* | 913 (61.2) |
|  | Other or no qualifications | 567 (38.0) |
|  | Don't know | 5 (0.3) |
|  | Prefer not to say | 8 (0.5) |
| Region where respondent lives; *n* (%) | England | 1258 (84.3) |
|  | Northern Ireland | 26 (1.7) |
|  | Scotland | 135 (9.0) |
|  | Wales | 74 (5.0) |
| Relationship status; *n* (%) | Married/living with partner | 910 (61.1) |
|  | In relationship but not living together | 126 (8.4) |
|  | Single | 337 (22.6) |
|  | Widowed | 14 (0.9) |
|  | Divorced | 71 (4.8) |
|  | Separated | 22 (1.5) |
|  | Prefer not to say | 12 (0.8) |
|  | Missing | 1 (0.1) |

Percentages may not sum to 100 owing to rounding. SD = standard deviation.
\*Undergraduate (e.g. BA, BSc) or postgraduate (e.g. MA, MSc, PhD) degree or other technical, professional or higher qualification.

### Cervical screening practice

Participation in cervical screening and associated factors are presented in Table 2.

**Table 2.**
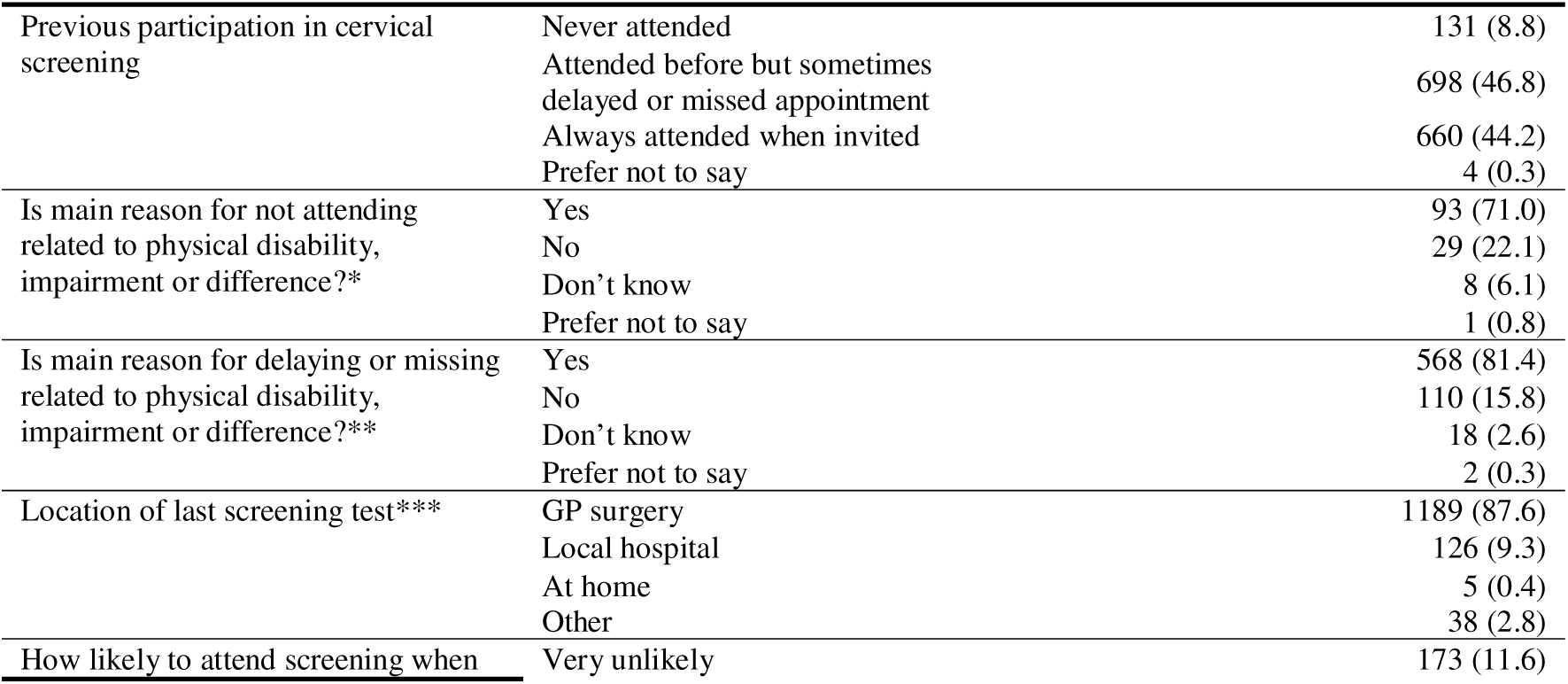
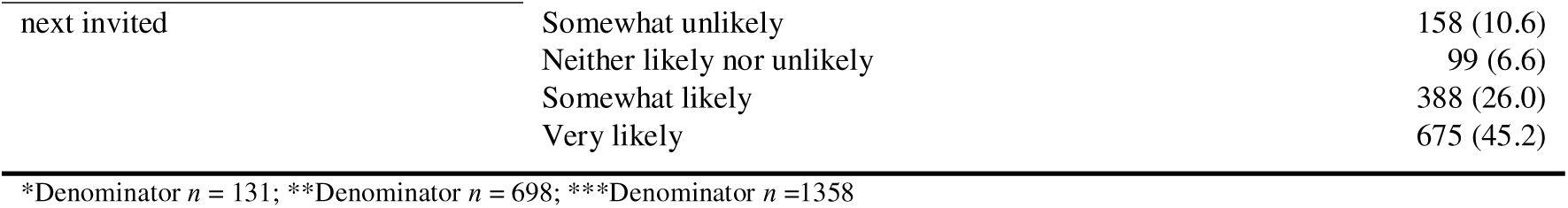
Cervical screening practices, *n* (%) (denominator *n* = 1493 unless otherwise indicated)

### Physical conditions

Table 3 presents the distribution of reported areas affected by physical condition among 1,493 participants. Most commonly reported were pain (67.2%) and mobility (58.9).

**Table 3.**
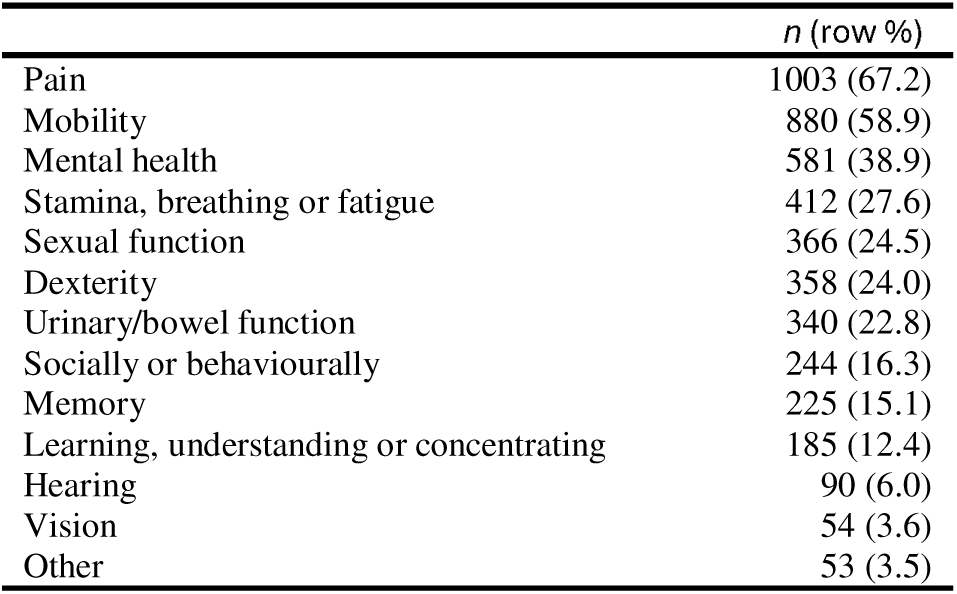
Areas affected by physical condition. Participants could endorse more than one area.

|  | *n* (row %) |
| --- | --- |
| Pain | 1003 (67.2) |
| Mobility | 880 (58.9) |
| Mental health | 581 (38.9) |
| Stamina, breathing or fatigue | 412 (27.6) |
| Sexual function | 366 (24.5) |
| Dexterity | 358 (24.0) |
| Urinary/bowel function | 340 (22.8) |
| Socially or behaviourally | 244 (16.3) |
| Memory | 225 (15.1) |
| Learning, understanding or concentrating | 185 (12.4) |
| Hearing | 90 (6.0) |
| Vision | 54 (3.6) |
| Other | 53 (3.5) |

The optional open-ended question ‘Please can you tell us the nature of your physical disability, condition, impairment or difference’ was answered by 1337 respondents; Table 4 shows the most frequently described conditions, some of which affect the body generally while others are specific to the pelvic region.

**Table 4.**
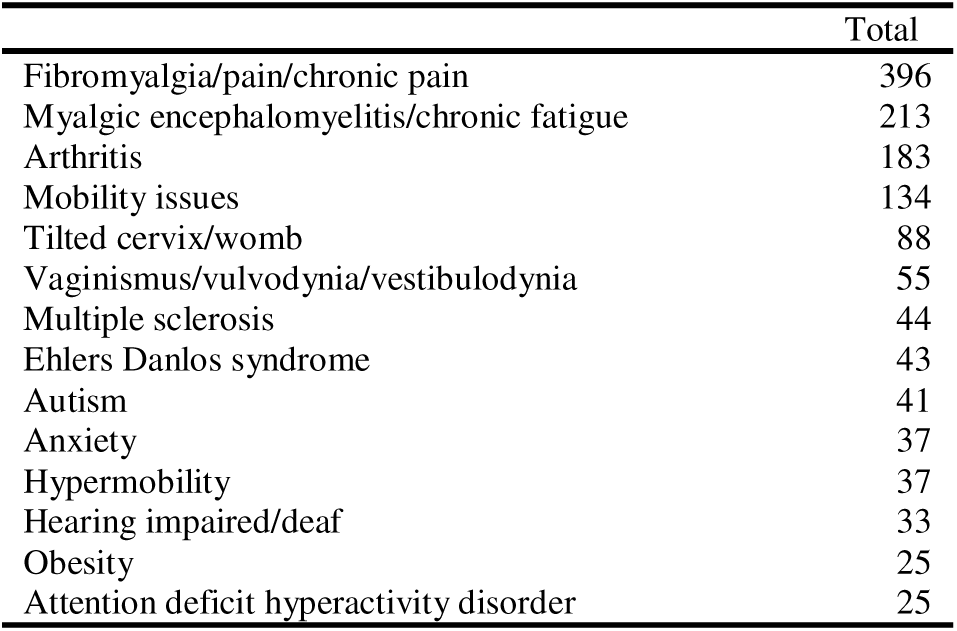
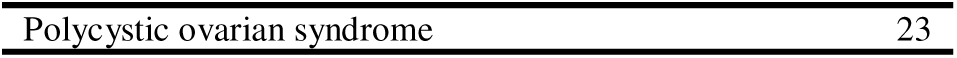
Most frequent responses describing physical disability, condition, impairment or difference (*n*=1337). Participants could nominate more than one condition.

As shown in Table 5, participants reported a range of assistive devices used to support daily functioning.

**Table 5.** Assistive devices used; participants could endorse more than one device, *n* = 1493.

|  | *n* (row %) |
| --- | --- |
| Walking stick | 240 (16.1) |
| Wheelchair | 156 (10.4) |
| Glasses | 127 (8.5) |
| Crutches | 107 (7.2) |
| Hearing aid | 57 (3.8) |
| Orthotic device | 56 (3.8) |
| Walking frame | 43 (2.9) |
| Magnifier | 11 (0.7) |
| White cane | 9 (0.6) |
| Communication board | 2 (0.1) |
| Hand-powered tricycle | 1 (0.1) |
| Artificial limb(s) | 1 (0.1) |
| Other | 39 (2.6) |

### Problems and solutions

The highest level of agreement (‘strongly agree’ and ‘somewhat agree’ combined) for problems related to screening (top ten shown in Table 6) was for concerns about pain (83.3%). Other items elicited at least 47.6% agreement (a list of all 36 problems provided is in supplementary materials Table A1).

**Table 6.** Attitudes and beliefs related to the problems with cervical screening (the 10 items eliciting the greatest agreement), *n* (%). Denominator *n* = 1493.

| **Statement** | **Strongly disagree** | **Somewhat disagree** | **Neither agree nor disagree** | **Somewhat agree** | **Strongly agree** | **% somewhat or strongly agree** |
| --- | --- | --- | --- | --- | --- | --- |
| I worry that a screening test will be painful | 70 (4.7) | 99 (6.6) | 79 (5.3) | 565 (37.8) | 680 (45.5) | 83.3 |
| Cervical screening tests are embarrassing | 143 (9.6) | 192 (12.9) | 106 (7.1) | 694 (46.5) | 358 (24.0) | 70.5 |
| I'm scared of what a screening test might find | 99 (6.6) | 149 (10.0) | 216 (14.5) | 663 (44.4) | 366 (24.5) | 68.9 |
| The examination table is uncomfortable | 182 (12.2) | 145 (9.7) | 166 (11.1) | 642 (43.0) | 358 (24.0) | 67.0 |
| I've had a bad experience of screening test in past | 194 (13.0) | 182 (12.2) | 145 (9.7) | 480 (32.2) | 492 (33.0) | 65.2 |
| Having to lie on my back with my legs apart is a problem | 249 (16.7) | 170 (11.4) | 147 (9.8) | 507 (34.0) | 420 (28.1) | 62.1 |
| I worry about having a male doctor or a male nurse do the test | 257 (17.2) | 200 (13.4) | 143 (9.6) | 412 (27.6) | 481 (32.2) | 59.8 |
| I intend to go when due, but don't always get around to is straight away | 345 (23.1) | 265 (17.7) | 116 (7.8) | 559 (37.4) | 208 (13.9) | 51.3 |
| Parking near to the GP surgery is limited/unavailable | 433 (29.0) | 216 (14.5) | 120 (8.0) | 431 (28.9) | 293 (19.8) | 48.7 |
| Doctors and nurses lack understanding of disability | 207 (13.9) | 302 (20.2) | 274 (18.4) | 504 (33.8) | 206 (13.8) | 47.6 |

The most endorsed potential solution to make cervical screening easier (‘strongly agree’ and ‘somewhat agree’ combined; the top ten shown in Table 7) was having a doctor or nurse who is willing to try different solutions (89.1%). Other solutions elicited at least 62.3% agreement (a list of all 19 solutions provided is in supplementary materials Table A2).

**Table 7.** Things that would or do make cervical screening easier (the 10 items eliciting the greatest agreement), *n* (%). Denominator *n* = 1493 except where indicated otherwise.

| Statement | Strongly disagree | Somewhat disagree | Neither agree nor disagree | Somewhat agree | Strongly agree | % somewhat or strongly agree |
| --- | --- | --- | --- | --- | --- | --- |
| A doctor or nurse who is willing to try different solutions | 31 (2.1) | 21 (1.4) | 111 (7.4) | 529 (35.4) | 801 (53.7) | 89.1 |
| A doctor or nurse who discusses my specific needs | 42 (2.8) | 27 (1.8) | 118 (7.9) | 587 (39.3) | 719 (48.2) | 87.5 |
| A doctor or nurse who understands my physical disability | 55 (3.7) | 38 (2.5) | 164 (11.0) | 593 (39.7) | 643 (43.1) | 82.8 |
| A smaller speculum | 55 (3.7) | 52 (3.5) | 200 (13.4) | 407 (27.3) | 779 (52.2) | 79.5 |
| A cervical screening passport (e.g. description of needs and what has/has not worked before) | 97 (6.5) | 72 (4.8) | 223 (14.9) | 554 (37.1) | 547 (36.6) | 73.7 |
| An examination couch that can be lowered | 201 (13.5) | 57 (3.8) | 187 (12.5) | 471 (31.5) | 577 (38.6) | 70.1 |
| Being invited by email or text | 88 (5.9) | 65 (4.4) | 298 (20.0) | 563 (37.7) | 479 (32.1) | 69.8 |
| Having a conversation with the doctor or nurse before the appointment | 92 (6.2) | 142 (9.5) | 227 (15.2) | 611 (40.9) | 421 (28.2) | 69.1 |
| Being offered medication that prevents cramps and spasms or other medication before the appointment* | 149 (10.0) | 155 (10.4) | 258 (17.3) | 496 (33.3) | 433 (29.0) | 62.3 |
| Being offered a longer appointment* | 116 (7.8) | 160 (10.7) | 377 (25.3) | 503 (33.7) | 335 (22.5) | 67.2 |

\* 2 missing values

**Table 8.**
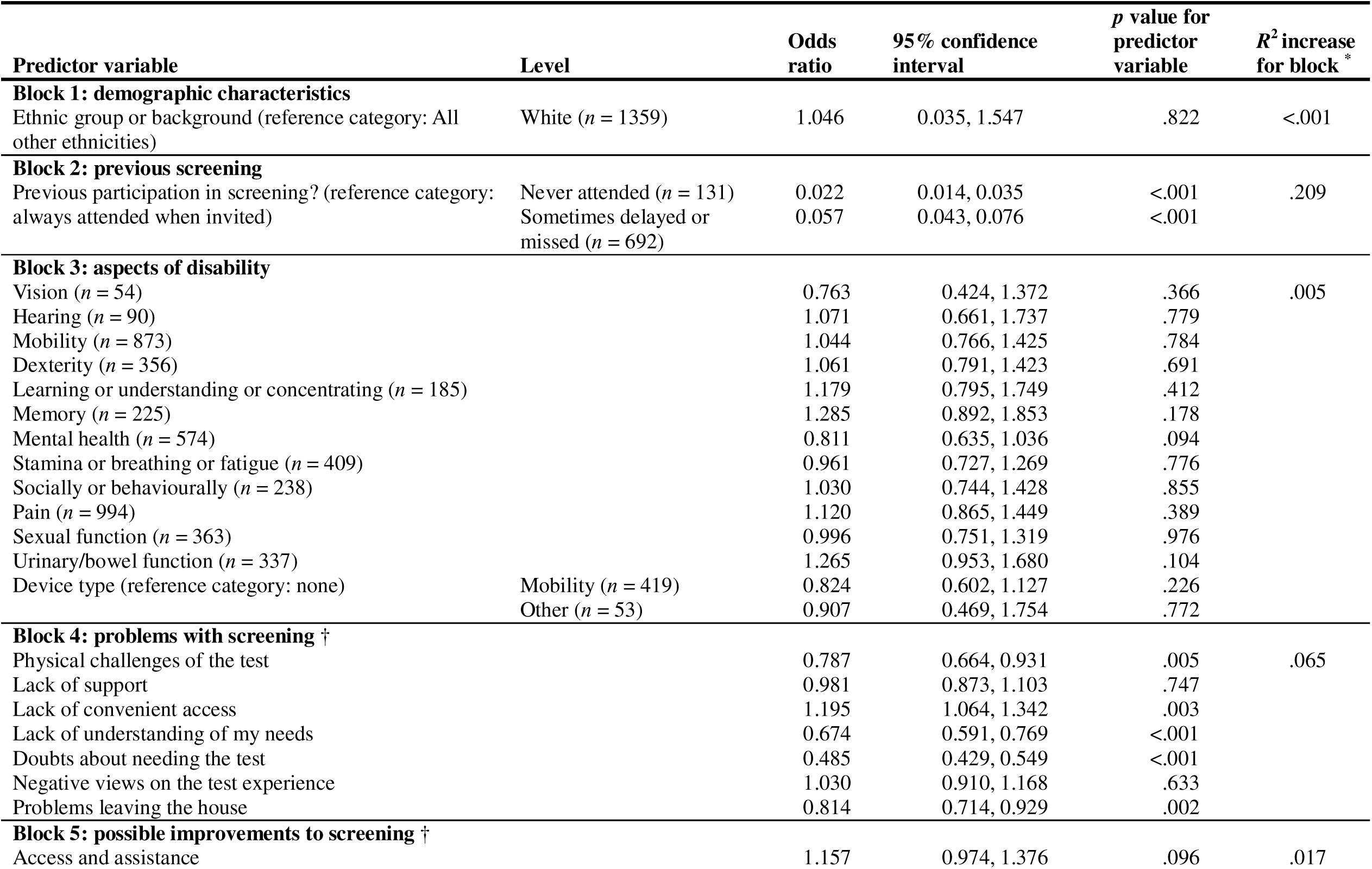
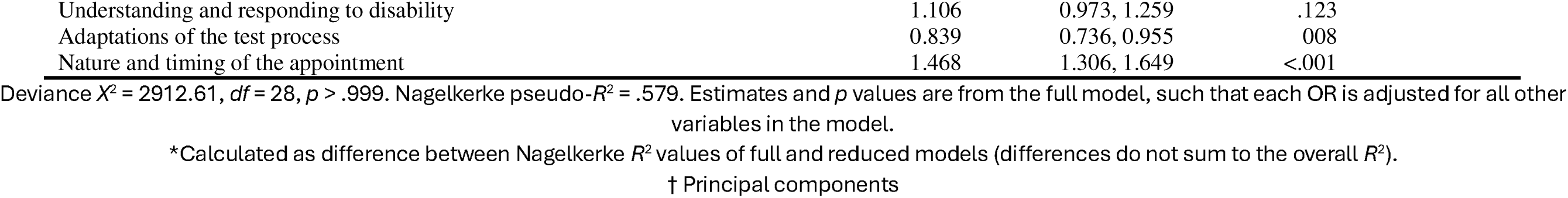
Ordinal logistic regression for screening preferences; *n* = 1480. The analysis excludes respondents who had missing values on one or more variables in the model (*n* = 13).

### Predictors of future screening attendance

The ordinal regression fitted the data well (*p* > .999) and explained 57.9% of the values on the outcome variable (Nagelkerke *R*^2^ = .579). Of the blocks in the model, previous screening participation (Block 2) had by far the greatest predictive power, explaining 20.9% of the outcome’s values. Compared to consistent previous attendance, previous never-attendance was a strong negative predictor of future screening attendance (OR = 0.022, 95% CI 0.014, 0.035; *p* < .001), as was previously delaying or missing appointments, though less strongly (OR = 0.057, 95% CI 0.043, 0.076; *p* < .001). In the remaining blocks in the model, the principal components relating to the problems with screening explained 6.5% of values, followed by the components relating to possible improvements in screening (1.7%). Among the problems with screening, four principal components negatively predicted future screening attendance, in increasing magnitude of association: ‘Problems leaving the house’ (OR 0.814, 95% CI 0.714, 0.929; *p* = .005), ‘Physical challenges of the test’ (OR 0.787, 95% CI 0.664, 0.931; *p* = .005) ‘Lack of understanding of my needs’ (OR 0.674, 95% CI 0.591, 0.769; *p* < .001) and ‘Doubts about needing the test’ (OR 0.485, 95% CI 0.429, 0.549; *p* < .001). Among components representing possible improvements to the test, significant predictors were: ‘Nature and timing of the appointment’ (OR 0.1.468, 95% CI 0.1.306, 1.469; *p* < .001) and ‘Adaptations of the test process’ (OR 0.839, 95% CI 0.736, 0.955; *p* = .008). Whilst the former positively predicted future screening attendance, the latter was a negative predictor. Neither ethnicity (Block 1) nor any of the aspects of disability (Block 3) were significant predictors.

### Qualitative findings

Six themes were identified (Table 9), representing participants’ experiences of cervical screening. Themes addressed the question of problems and solutions relating to engaging in cervical screening.

**Table 9.** Table of themes.

| Theme | Description |
| --- | --- |
| 1. Bodily and pain-related problems | The physical and bodily experience of cervical screening: problems of getting into the required position, pain experienced during screening, and how screening would trigger symptoms related to specific physical conditions. |
| 2. Anxiety and psychological distress | The psychological and emotional impact of screening, including anxiety about how the test will feel and what the test will find, feeling embarrassed and self-conscious during screening. |
| 3. Trauma from sexual abuse or healthcare experiences | The impact of having previous trauma or sexual abuse that impacts how participants feel about screening. |
| 4. Procedural problems affecting screening uptake | Procedural issues that might affect attendance, including organizational problems. These included stress of booking and attending appointments, need for additional support, and limited appointment availability. |
| 5. Equipment and physical environment as problematic | The structural and environmental problems that patients encounter when accessing screening. Difficulty accessing the GP surgery, examination room, examination table, lack of accessible transport and experiencing an uncomfortable environment when at the surgery. |
| 6. Healthcare professionals' lack of compassion, empathy and knowledge | The attitudinal issues faced during screening. Difficulty discussing own needs, healthcare professionals (HCPs) lacking empathy and understanding of disabilities, representing lack of information provision and communication. Including having to explain personal issues at every screening appointment and notes not being checked or shared between HCPs. |

#### Theme 1. Bodily and pain-related problems

Many participants stated that they found cervical screening painful. The pain was not just associated with speculum insertion but extended to the pain experienced when trying to manoeuvre into the required position for screening (lying on one’s back with legs apart). Participants described how screening would also trigger pain linked to their physical disability or condition:

> *The last time I attended my appointment at the Dr’s the nurse asked me to sit on the edge of the bed then to lie back on the bed. This was extremely painful due to my chronic back pain.* (Participant with multiple sclerosis)

Another participant described experiencing pain from speculum insertion and how this would last after the examination, with her ability to walk also affected by the procedure triggering hip pain:

> *I found the whole thing painful. I know it’s important and I should get it done, but I just can’t face it. I could hardly walk afterwards due to my hip and the pain of getting the speculum in and the scraping feeling was awful.* (Participant with fibromyalgia)

One participant described how she could not predict how her symptoms would be on the day of screening. She described not being able to tolerate screening on certain days due to the pain it would cause:

> *Pain. I suffer with a lot of vaginal/cervix pain due to scar tissue and on bad days no way could I consider having a smear. But you can’t plan your bad days when you book an appointment and if you cancel at short notice you’re made to feel irresponsible and wasting an appointment.* (Participant with bladder and bowel prolapse)

When asked what would make cervical screening easier, participants gave responses centred around measures to reduce the pain experienced during the screening examination. One participant emphasized the solutions presented in the survey items:

> *These have been mentioned above but I’d like to emphasise them, a smaller speculum for less pain, a nurse who knows about my condition and is patient, and medication beforehand that can help with the pain.* (Participant with adenomyosis)

#### Theme 2. Anxiety and psychological distress

Participants described the psychological and emotional impact of having a cervical screening examination. Many participants wrote about anxiety around how the test would feel, with some expressing a fear of pain, and others mentioning feeling worried about what the test would find. One participant wrote about how she felt unable to face having the test due to worry:

> *I really want to get tested, but I cannot face it. I may feel brave enough to make the appointment on a particular day, but I wouldn’t feel brave enough to go as I would worry about it and make myself really unwell.* (Participant with a mental health condition)

Anxiety regarding incontinence or bleeding was also a concern for several women. One woman who had found the screening process to be undignified, and the practice staff to be rude, said that she intended to use a home HPV test instead of attending the surgery.

Delaying a cervical screening test due to fear of what the test might find was mentioned by another participant: ‘*One of the main reasons I don’t like attending is fear of them detecting cervical cancer which sometimes results in me delaying attending a screening*’ (Participant with mobility issue).

To help reduce anxiety, participants emphasized two solutions presented in the survey: a cervical screening passport and HPV self-sampling. One woman wrote:

> *I hadn’t heard of a cervical screening passport before though, I really like the sound of that. I think it would help with the anxiety in the run up and would streamline the process for me. The anxiety and stress in the week before causes increased pain, fatigue and tension which all impacts getting the test done.* (Participant with fibromyalgia)

Participants described how HPV self-sampling tests could help to reduce the discomfort and embarrassment of the screening examination, and some women reported having previously used private self-sampling for similar reasons, one of whom pleaded for this to be made available on the NHS.

#### Theme 3. Trauma from sexual abuse or healthcare experiences

Participants wrote about their experience of trauma, and some described having experienced sexual abuse. One participant discussed how the cervical screening procedure could elicit memories of previous traumatic experiences of sexual abuse and affect her mental health:

> *Having been subjected to sexual abuse having cervical screening is harder for me mentally than physically. It leaves me feeling anxious, “dirty”, having flash backs and generally low for months after. I have never told anyone about it as it happened years ago so there is no point bringing it up now but having to face cervical screening is one of the worst triggers for me as I generally cope pretty well until that letter drops through the door then I spend months trying to build up the courage to go.* (Participant with degenerative disc disease)

Experiencing previous medical trauma was also discussed by participants. One participant described how, due to previous medical trauma, she finds it hard to seek medical help, which prevents her from accepting her invitation to cervical screening:

> *Previous medical trauma makes it very hard for me to seek medical help, even when I desperately need it. I almost died due to medical neglect in the past, and I feel that if they won’t help in an acute life and death situation, then why would I accept a screening, when I’m asymptomatic, and my debilitating issues have been deemed not worthy of help.* (Participant with fibromyalgia/chronic fatigue syndrome)

For some participants who had experienced previous sexual abuse or trauma being able to use an HPV self-sampling kit would help them to feel more at ease to complete their screening tests:

> *I would never want to go into the surgery and have someone do this, in part because of my history of sexual abuse and lack of treatment for PTSD, and in part because of my disability and being housebound (and struggling with GPs or nurses understanding this). I would, however, be completely comfortable doing it myself at home. That would be a game-changer.* (Participant with mobility issues)

#### Participants also emphasized the importance of being reassured that a female healthcare professional would perform their cervical screening test.Theme 4. Procedural problems affect screening uptake

This theme described the problems participants faced in attending the screening appointment, which included limited appointment availability causing stress when booking appointments and not having enough time during the procedure due to having additional needs. One participant described the impact of limited appointment availability:

> *Appointments are only offered in one fixed slot on a particular day of the week by my GP surgery. This makes attending really difficult as I may have to change work commitments or not have care support available*. (Participant with a spinal cord injury)

A few participants found that limited availability of appointments made it hard to attend when not menstruating and another had found that a flare-up of her condition had prevented her from attending her last appointment. One woman felt able to attend when having a rare ‘good day’, but these days are unpredictable, and she cannot get a same-day appointment to accommodate them.

When describing what would make screening easier, one participant indicated that longer appointments would help to allow time for her to explain to the practitioner the difficulties she faces. Another woman noted that feeling that she was listened to and not being rushed would make a huge difference to her experience of screening.

Several participants also discussed the idea of a walk-in clinic to reduce the need to book an appointment in advance. They described how this would ensure that they could attend appointments at suitable times when they could receive the additional help and support to suit their needs. This would support participants with conditions that cause unpredictable symptoms and make booking appointments in advance difficult.

> *A walk-in clinic where you don’t have to make a prior appointment – if your physical condition is very variable and changes day to day or hour to hour then it is hard to book advance appointments. A walk-in clinic would be so much more accessible as this stops you being made to feel bad or guilty if you have to cancel an appointment at the last minute and means you don’t get a black mark against you.* (Participant with autoimmune disease)

#### Theme 5. Equipment and physical environment as problematic

Participants discussed how they faced problems during their screening appointments due to a lack of equipment to suit their needs. They described difficulty getting onto the examination table, which they sometimes considered too high and/or lacking a grab rail, and a lack of support to assist them. One participant described how her disability requires assistance or supportive aids; however, this was unavailable at her GP practice:

> *My disability prevents me from being able to stand up from my wheelchair and carers do not lift. There is no hoist or bed that can be lowered or raised at my doctors surgery so this makes it extremely difficult for me to have a smear test done at my doctors surgery. I am also unable to climb up any step or stool to get onto the bed.* (Participant with muscular dystrophy)

Alongside inaccessible examination tables, other equipment such as speculums is also an issue for some participants. One participant described needing a different-sized speculum and experiencing bleeding:

> *The equipment they use, the speculum. I have a long vagina but the different sizes make things more uncomfortable, opening wider. I find the whole process really painful and the nurses really struggle to get the samples they need. In the end I bleed and that affects the results.* (Participant with a tilted uterus)

The physical environment in the examination room was also problematic for several participants. One participant discussed that the environment in the room can make her feel vulnerable and stated that a privacy curtain could help:

> *I think the way the rooms are set up has an impact on women. A room that is very open with bright lights and open spaces can feel vulnerable to be undressed etc. I think at least a curtain should be drawn around beds to offer privacy in case the door gets opened.* (Participant with Ehlers Danlos syndrome)

#### Theme 6. HCPs’ lack of compassion, empathy and knowledge

This theme described the participants’ experiences with the HCPs they have contact with during the screening process. Often, participants wrote about feeling a lack of empathy and poor understanding of disabilities from their HCPs. One participant wrote about a HCP demonstrating a lack of understanding and ignorance about her disorder, which affected her emotional wellbeing:

> *More ignorance of my disorder. I have a memory of going for a smear, explaining what happens when I have a smear with a lot of bleeding. She not only gasped when she was doing the smear, she said that I don’t just bleed I “pour”. I sat in the car crying for over 30 mins after and could not drive until my legs stopped shaking. It makes it so hard to go back.* (Participant with polycystic ovarian syndrome)

Another participant also wrote about a lack of empathy from HCPs relating to the pain she experiences during cervical screening. Some participants reported that HCPs lacked understanding of, or did not believe, their pain and were told that a screening examination should not be painful:

> *Medical professionals not taking the time/caring enough about pudendal neuralgia, so when I say it will hurt me they just say “oh it shouldn’t it’s just a smear” – I’m always frustrated because I’m trying to express that my disability means it will be painful for me.* (Participant with pudendal neuralgia)

One woman suggested that the failure to provide sedation was related to HCP lack of understanding of vaginismus.

Another woman described how she found the screening process a negative experience both physically and psychologically:

> *I hate my local nurses doing the NHS smear test. They have an attitude of ‘pull your big girl pants up and get on with it’. Like something out of the Victorian era. The whole process is demeaning, embarrassing and very uncomfortable… I have raised my concerns to the practice manager politely and respectfully, but they don’t care. In their eyes I’m just a winger [whinger] who needs to go away and so I am ignored.* (Participant with Ehlers Danlos syndrome)

For one woman, a lack of understanding and compassion from staff had caused her to move to another practice.

Participants also mentioned how they felt their medical notes were not checked by HCPs prior to their screening appointments and this meant having to explain their physical condition or needs at every appointment:

> *I always have to tell them that I have a bladder prolapse so they don’t get a shock. They’ve never known before even though it’s in my notes. There could be medical conditions at the top of every patient’s notes so that doctors and nurses see a snapshot before you come in and can adapt their appointment.* (Participant with osteoarthritis)

In response to what would make cervical screening easier, many participants stated that more empathetic HCPs would help to improve patient experience. A greater understanding of physical disabilities from knowledgeable HCPs would help patients feel listened to at the appointment and could ensure that individual needs are met:

> *Having the cervical screening invite team KNOW I’m wheelchair and hoist dependent and send information that’s relevant to my needs and location. My GP had no idea what to do with me and I’ve been left without a test as a result.* (Wheelchair user)

Better staff training focused on specific physical conditions and how to deal with patients with additional needs was also discussed by participants:

> *Knowing that the nurses will treat you with respect and understanding and allowed you to voice your concerns without them getting annoyed. Having nurses that are trained specifically to deal with patients with endometriosis would fill be more confidence. And if they could take it seriously when we say we are in pain.* (Participant with endometriosis)

## Discussion

While a large minority of our sample had always attended cervical screening when invited, for those who had delayed, missed or never attended screening, the majority reported disability-related factors to be the main reason. The problems that most participants endorsed fell into two broad categories: 1) problems faced by non-disabled women such as concerns about pain and embarrassment and 2) problems that were specific to disability such as the examination table being uncomfortable and it being challenging to get into the correct position on the couch. While it is unsurprising that Disabled women face the same problems as non-disabled women, it is noteworthy the extent to which the Disabled women in our sample agreed with those statements. In our survey, 83.3% were worried that a screening test would be painful and 70.5% agreed cervical screening tests are embarrassing. This contrasts with 14% and 29% respectively of a general UK sample endorsing the same items (Waller et al 2009) and 38% and 21% respectively in a more recent UK sample (Whitelock et al, 2024) with the same questions phrased slightly differently. Most of the open-ended comments fell into the disability-related category, and these barriers are consistent with the findings from a qualitative study of 56 women in the United States which reported that accessibility barriers and clinician ableism led some participants to delay or avoid screening (Vinson et al, 2025). We found that previous screening attendance predicted future screening attendance. Additionally, problems related to leaving the house, the physical challenges of the test, lack of understanding of patient needs and doubts about needing the test all negatively predicted future screening attendance. Considering the wide range of problems, it is important that delaying or missing a screening appointment is not simply interpreted as non-compliance or disengagement.

In terms of potential solutions, the most frequently endorsed items were having a doctor or nurse who is willing to try different solutions, discusses specific needs, and understands physical disability. The need for this was powerfully underscored by our thematic analysis, particularly theme six, which was concerned with healthcare professionals’ lack of compassion, empathy and knowledge. For many of our participants, it is these attitudinal issues rather than the condition itself which is the problem. Other solutions related to speculum size, the examination couch, appointment time and opportunity for discussion before the appointment, and a cervical screening passport that would facilitate the HCP interactions. One of the solutions, being invited by email or text, has since been addressed by the introduction of a digital messaging platform in England (NHSNotify: https://digital.nhs.uk/about-nhs-digital/digital-first-messaging/transforming-cervical-screening-communications). Solutions also influenced future screening intention, with those who endorsed an improvement in the nature and timing of the test process being more likely to attend screening and those who wanted adaptations to the test procedure being less likely to attend. Since participants were asked about solutions that would, or already do, make screening more accessible, it may be that this reflects the fact that the nature and timing of the test is already flexible for many women so they plan to attend as this is accommodated, while adaptations are not yet so widely in place and thus this might deter women.

In the open-ended comments, some women reported being keen to engage with HPV self-sampling and this was also reflected in the survey questions on HPV self-sampling reported by Kemp et al (2026). It is certainly likely to be the case that for many Disabled women the availability of vaginal self-sampling will make cervical screening more accessible – or does so, for those countries that have already introduced it. There are also other types of home testing methods such as urine sampling in development (Davies et al, 2025). However, women in the UK who test positive for HPV will need to access speculum-based cervical screening for follow-up tests; so these tests should not be seen as an alternative to making conventional screening more accessible.

### Strengths and limitations

We recruited a large sample of women who self-identified as having a physical disability, condition, impairment or difference. This wording was informed by our PPI work and the social model of disability to be maximally inclusive of conditions that fluctuate, are invisible, or may not be captured by medicalized definitions of disability. Our PPI work also enabled us to include questions in our survey to capture problems affecting Disabled women that have been overlooked in previous research. If the problems faced by Disabled people are not captured because they are not asked about, policy and practice cannot evolve to address them, and health inequities will persist.

Limitations of the research include that cervical screening status was self-reported, that we only recruited women who found cervical screening difficult or impossible to attend, so the findings may not reflect the experiences of other Disabled women, and that we did not over-sample for intersecting characteristics such as ethnicity to enable us to explore intersecting problems and solutions.

### Implications for clinical practice

Current advice about how to support women with learning disabilities in the UK (Gov.uk, 2026), such as a preliminary visit to the practice or longer appointment time, could usefully be extended to physically Disabled women. While physical adjustments such as height-adjustable couches and accessible spaces may not be feasible for all surgeries due to space or cost restrictions; these could be made available in women’s health hubs or in selected practices in each area so that women can be referred there. Most importantly, it is crucial that HCPs receive training in how best to support physically Disabled women to ensure that the woman’s experience of her own condition and of cervical screening is respected and that she is supported to have the most dignified and comfortable procedure possible.

## Conclusion

This is the first survey in the UK to explore problems and solutions related to cervical screening from the perspective of physically Disabled women. It was developed with extensive PPI involvement and using the lens of the social model of disability to ensure that the perspective and needs of women were centred. Physically Disabled women experience a wide range of problems related to cervical screening, with additional problems to those faced by non-disabled women. Disability-specific problems need particular attention, most pressing of which is the need for disability-informed training for sample-takers to reduce health inequities for this population.

## Data Availability

The quantitative data are available at: https://osf.io/ufx8r/files/tm2us
The qualitative data will be available from the OSF project page following peer review: https://osf.io/ufx8r/overview

## Acknowledgements

We would like to thank the following members of our PPI stakeholder group for their work in helping us develop the survey materials: Susan Bennett, Victoria Garcia, Lynda Hesketh BEM, Alycia Hirani, Emma Major, Georgina Moore, Alison Murray, Melody Powell, Iqra Saeed, Roxanne Steel, plus a further four members who prefer to remain anonymous. We would also like to thank the people who took part in the survey, as well as SCOPE, Spinal Injuries Association, Brittle Bone Society, and Muscular Dystrophy UK for circulating the survey, and Jo’s Cervical Cancer Trust and The Eve Appeal for providing support to this project.

## Declaration of competing interests

Susan Sherman is a Trustee for the British Society for Colposcopy and Cervical Pathology. The authors declare no other potential competing interests with respect to the research, authorship, and/or publication of this article.

## Funding

This work is funded by the UK National Institute for Health and Care Research (NIHR) under its Research for Patient Benefit (RfPB) Programme (Grant Reference Number NIHR204322). The views expressed are those of the authors and not necessarily those of the NIHR or the Department of Health and Social Care. CCG is part funded by the National Institute for Health and Care Research (NIHR) Applied Research Collaboration West Midlands (NIHR200165).

## Ethical considerations

This study was granted ethical approval from Keele University Research Ethics Committee (REF: REC Project Reference 0787) and the University of Sheffield School of Psychology research ethics committee (REF: 064385).

## Footnotes

[1] A note on terminology. This research has been informed by extensive stakeholder engagement, and we are using the wording preferred by our stakeholders to reflect the Social Model of Disability whereby disability is created by the way the environment is structured rather than by a specific condition or impairment: hence a Disabled person rather than a person with a disability. The capitalization of Disabled acknowledges a collective cultural identity and emphasizes the term’s political significance (Disability Rights UK, 2026).

