## Supplementary Material for "Understanding problems and solutions related to accessing cervical screening for people with a physical disability, condition, impairment or difference"

**SUPPLEMENTARY MATERIALS**

**Table A1.** Reponses to items relating to problems with cervical screening:

| **Please indicate how much you agree with each of the following statements about cervical screening** | **Strongly disagree** | **Disagree** | **Neither agree nor disagree** | **Agree** | **Strongly agree** | **Missing** |
| --- | --- | --- | --- | --- | --- | --- |
| Cervical screening tests are embarrassing | 143 (9.6) | 192 (12.9) | 106 (7.1) | 694 (46.5) | 358 (24.0) | 0 (0.0) |
| I’ve had a bad experience of a cervical screening test in the past | 194 (13.0) | 182 (12.2) | 145 (9.7) | 480 (32.2) | 492 (33.0) | 0 (0.0) |
| I do not need a test if I do not have any symptoms | 1093 (73.2) | 285 (19.1) | 66 (4.4) | 40 (2.7) | 9 (0.6) | 0 (0.0) |
| I worry that a cervical screening test will be painful | 70 (4.7) | 99 (6.6) | 79 (5.3) | 565 (37.8) | 680 (45.5) | 0 (0.0) |
| I’m scared of what a cervical screening test might find | 99 (6.6) | 149 (10.0) | 216 (14.5) | 663 (44.4) | 366 (24.5) | 0 (0.0) |
| I do not trust the cervical screening test | 829 (55.5) | 427 (28.6) | 149 (10.0) | 68 (4.6) | 20 (1.3) | 0 (0.0) |
| I don't feel at risk of cervical cancer | 494 (33.1) | 414 (27.7) | 389 (26.1) | 163 (10.9) | 33 (2.2) | 0 (0.0) |
| I am immunocompromised/at higher risk of infections | 602 (40.3) | 267 (17.9) | 184 (12.3) | 249 (16.7) | 191 (12.8) | 0 (0.0) |
| I have a history of sexual abuse | 863 (57.8) | 137 (9.2) | 104 (7.0) | 210 (14.1) | 179 (12.0) | 0 (0.0) |
| I’m not sexually active so I don’t need to go for a cervical screening test | 956 (64.0) | 273 (18.3) | 134 (9.0) | 92 (6.2) | 38 (2.5) | 0 (0.0) |
| Doctors and nurses assume I am not sexually active | 701 (47.0) | 248 (16.6) | 372 (24.9) | 116 (7.8) | 56 (3.8) | 0 (0.0) |
| I intend to go when I am due, but I don’t always get round to it straight away | 345 (23.1) | 265 (17.7) | 116 (7.8) | 559 (37.4) | 208 (13.9) | 0 (0.0) |
| It is difficult to get an appointment to fit in with work/childcare commitments | 375 (25.1) | 265 (17.7) | 202 (13.5) | 420 (28.1) | 232 (15.5) | 0 (0.0) |
| There are additional costs associated with attending screening | 774 (51.8) | 329 (22.0) | 123 (8.2) | 193 (12.9) | 74 (5.0) | 0 (0.0) |
| I cannot leave the house | 923 (61.8) | 288 (19.3) | 107 (7.2) | 149 (10.0) | 26 (1.7) | 0 (0.0) |
| I don't have enough support from family/friends | 791 (53.0) | 312 (20.9) | 165 (11.1) | 172 (11.5) | 53 (3.5) | 0 (0.0) |
| I don’t have the right support from my personal assistant/carer/support worker | 861 (57.7) | 159 (10.6) | 389 (26.1) | 60 (4.0) | 24 (1.6) | 0 (0.0) |
| I don’t have a care and support plan (sometimes called direct payment) | 609 (40.8) | 102 (6.8) | 254 (17.0) | 104 (7.0) | 424 (28.4) | 0 (0.0) |
| I don’t have enough hours in my care and support plan | 772 (51.7) | 108 (7.2) | 532 (35.6) | 43 (2.9) | 38 (2.5) | 0 (0.0) |
| Getting transport to the appointment is hard or impossible | 723 (48.4) | 289 (19.4) | 146 (9.8) | 280 (18.8) | 55 (3.7) | 0 (0.0) |
| My GP surgery is not in a convenient location | 660 (44.2) | 379 (25.4) | 112 (7.5) | 259 (17.3) | 83 (5.6) | 0 (0.0) |
| Parking near to the GP surgery is limited/unavailable | 433 (29.0) | 216 (14.5) | 120 (8.0) | 431 (28.9) | 293 (19.6) | 0 (0.0) |
| Getting into the surgery is difficult | 591 (39.6) | 324 (21.7) | 145 (9.7) | 321 (21.5) | 112 (7.5) | 0 (0.0) |
| The examination room can't accommodate my mobility aid | 881 (59.0) | 223 (14.9) | 282 (18.9) | 74 (5.0) | 33 (2.2) | 0 (0.0) |
| I'm unable to undress myself | 957 (64.1) | 256 (17.1) | 83 (5.6) | 151 (10.1) | 46 (3.1) | 0 (0.0) |
| There is a lack of transfer equipment (e.g. hoist) | 735 (49.2) | 121 (8.1) | 421 (28.2) | 127 (8.5) | 89 (6.0) | 0 (0.0) |
| Getting onto the examination table is difficult | 523 (35.0) | 171 (11.5) | 176 (11.8) | 430 (28.8) | 193 (12.9) | 0 (0.0) |
| Staying on the examination table is difficult | 597 (40.0) | 202 (13.5) | 196 (13.3) | 347 (23.2) | 151 (10.1) | 0 (0.0) |
| The examination table is uncomfortable | 182 (12.2) | 145 (9.7) | 166 (11.1) | 642 (43.0) | 358 (24.0) | 0 (0.0) |
| Having to lie on my back with my legs apart is a problem | 249 (16.7) | 170 (11.4) | 147 (9.8) | 507 (34.0) | 420 (28.1) | 0 (0.0) |
| I worry about having a male doctor or male nurse do the test | 257 (17.2) | 200 (13.4) | 143 (9.6) | 412 (27.6) | 481 (32.2) | 0 (0.0) |
| Doctors and nurses lack understanding of disability | 207 (13.9) | 302 (20.2) | 274 (18.4) | 504 (33.8) | 206 (13.8) | 0 (0.0) |
| Doctors and nurses are not willing to listen to me about my disability | 279 (18.7) | 376 (25.2) | 309 (20.7) | 386 (25.9) | 143 (9.6) | 0 (0.0) |
| Screening appointments are too short | 355 (23.8) | 352 (23.6) | 399 (26.7) | 297 (19.9) | 90 (6.0) | 0 (0.0) |

**Table A2.** Reponses to items relating to possible improvements to cervical screening:

| **Please indicate how much you agree that each item below would (or already does) make cervical screening easier for you to attend** | **Strongly disagree** | **Disagree** | **Neither agree nor disagree** | **Agree** | **Strongly agree** | **Missing** |
| --- | --- | --- | --- | --- | --- | --- |
| Being invited via email or text | 88 (5.9) | 65 (4.4) | 298 (20.0) | 563 (37.7) | 479 (32.1) | 0 (0.0) |
| Being offered a longer appointment | 116 (7.8) | 160 (10.7) | 377 (25.3) | 5.3 (33.7) | 335 (22.4) | 2 (0.1) |
| Having a conversation with the doctor or nurse before the appointment | 92 (6.2) | 142 (9.5) | 227 (15.2) | 611 (40.9) | 421 (28.2) | 0 (0.0) |
| Being offered medication that prevents muscle cramps and spasms (antispasmodic) or other medication before the appointment | 149 (10.0) | 155 (10.4) | 258 (17.3) | 496 (33.2) | 433 (29.0) | 2 (0.1) |
| Being offered a test at a different venue e.g. hospital or sexual health venue | 311 (20.8) | 279 (18.5) | 369 (24.7) | 316 (21.2) | 221 (14.8) | 0 (0.0) |
| A home visit from a doctor or nurse | 435 (29.1) | 278 (18.6) | 244 (16.3) | 267 (17.9) | 269 (18.0) | 0 (0.0) |
| Being provided with an ambulance or other transport to the appointment | 772 (15.7) | 241 (16.1) | 270 (18.1) | 155 (10.4) | 55 (3.7) | 0 (0.0) |
| A doctor or nurse who understands my physical disability | 55 (3.7) | 38 (2.5) | 164 (11.0) | 593 (39.7) | 643 (43.1) | 0 (0.0) |
| A doctor or nurse who discusses my specific needs | 42 (2.8) | 27 (1.8) | 118 (7.9) | 587 (39.3) | 719 (48.2) | 0 (0.0) |
| A cervical screening passport (e.g. a description of needs and what has worked/not worked before) | 97 (6.5) | 72 (4.8) | 223 (14.9) | 554 (37.1) | 547 (36.6) | 0 (0.0) |
| Wheelchair access to the surgery | 529 (35.4) | 116 (7.8) | 484 (32.4) | 130 (8.7) | 234 (15.7) | 0 (0.0) |
| Assistance with undressing/dressing | 608 (40.7) | 191 (12.8) | 332 (22.2) | 233 (15.6) | 129 (8.6) | 0 (0.0) |
| A hoist to get me on the bed | 731 (49.0) | 196 (13.1) | 387 (25.9) | 109 (7.3) | 70 (4.7) | 0 (0.0) |
| An examination couch that can be lowered | 201 (13.5) | 57 (3.8) | 187 (12.5) | 471 (31.5) | 577 (38.6) | 0 (0.0) |
| A sliding board/transfer board/pat slide to get me on the bed | 566 (37.9) | 186 (12.5) | 448 (30.0) | 167 (11.2) | 126 (8.4) | 0 (0.0) |
| A smaller speculum | 55 (3.7) | 52 (3.5) | 200 (13.4) | 407 (27.3) | 779 (52.2) | 0 (0.0) |
| Being allowed to insert the speculum myself | 381 (25.5) | 273 (18.3) | 367 (24.6) | 247 (16.5) | 224 (15.0) | 1 (0.1) |
| A doctor or nurse who is willing to try different solutions | 31 (2.1) | 21 (1.4) | 111 (7.4) | 529 (35.4) | 801 (53.7) | 0 (0.0) |
| Being offered the left lateral position for the screening test | 148 (9.9) | 111 (7.4) | 408 (27.3) | 463 (31.0) | 363 (24.3) | 0 (0.0) |

**Table A3.** Principal components relating to problems with screening. Data are component loadings; loadings less than .440 are not shown. The seven components explained 51% of the variance in the original items.

|  | Physical challenges of the test | Lack of support | Lack of convenient access | Lack of understanding of my needs | Doubts about needing the test | Negative views on the test experience | Problems regarding leaving the house to attend the test |
| --- | --- | --- | --- | --- | --- | --- | --- |
| Cervical screening tests are embarrassing |  |  |  |  |  | .622 |  |
| I’ve had a bad experience of a cervical screening test in the past |  |  |  |  |  | .409 |  |
| I do not need a test if I do not have any symptoms |  |  |  |  | .719 |  |  |
| I worry that a cervical screening test will be painful |  |  |  |  |  | .547 |  |
| I’m scared of what a cervical screening test might find |  |  |  |  |  | .563 |  |
| I do not trust the cervical screening test |  |  |  |  | .504 |  |  |
| I don't feel at risk of cervical cancer |  |  |  |  | .701 |  |  |
| I am immunocompromised/at higher risk of infections |  |  |  |  |  |  | .503 |
| I have a history of sexual abuse |  |  |  |  |  |  |  |
| I’m not sexually active so I don’t need to go for a cervical screening test |  |  |  |  | .741 |  |  |
| Doctors and nurses assume I am not sexually active |  |  |  |  |  |  |  |
| I intend to go when I am due, but I don’t always get round to it straight away |  |  |  |  |  |  |  |
| It is difficult to get an appointment to fit in with work/childcare commitments. |  |  | .602 |  |  |  |  |
| There are additional costs associated with attending screening |  |  | .464 |  |  |  |  |
| I cannot leave the house |  |  |  |  |  |  | .594 |
| I don't have enough support from family/friends |  | .548 |  |  |  |  |  |
| I don’t have the right support from my personal assistant/carer/support worker |  | .705 |  |  |  |  |  |
| I don’t have a care and support plan |  | .724 |  |  |  |  |  |
| I don’t have enough hours in my care and support plan |  | .772 |  |  |  |  |  |
| Getting transport to the appointment is hard or impossible |  |  |  |  |  |  | .491 |
| My GP surgery is not in a convenient location |  |  | .687 |  |  |  |  |
| Parking near to the GP surgery is limited/unavailable |  |  | .688 |  |  |  |  |
| Getting into the surgery is difficult |  |  | .623 |  |  |  |  |
| The examination room can't accommodate my mobility aid | .416 |  |  |  |  |  |  |
| I'm unable to undress myself | .602 |  |  |  |  |  |  |
| There is a lack of transfer equipment | .593 | .434 |  |  |  |  |  |
| Getting onto the examination table is difficult | .836 |  |  |  |  |  |  |
| Staying on the examination table is difficult (e.g. no handrails, no sides) | .814 |  |  |  |  |  |  |
| The examination table is uncomfortable | .656 |  |  |  |  |  |  |
| Having to lie on my back with my legs apart is a problem | .705 |  |  |  |  |  |  |
| I worry about having a male doctor or male nurse do the test |  |  |  |  |  | .586 |  |
| Doctors and nurses lack understanding of disability |  |  |  | .811 |  |  |  |
| Doctors and nurses are not willing to listen to me about my disability |  |  |  | .814 |  |  |  |
| Screening appointments are too short |  |  |  | .469 |  |  |  |

**Table A4**. Principal components relating to potential improvements in screening. Data are component loadings; loadings less than .440 are not shown (the largest loading for ‘Being offered the left lateral position’ was .348, on ‘Adaptations of the test process’). The four components accounted for 55% of the variance in the original items.

|  | Access and assistance | Understanding and responding to disability | Adaptations of the test process | Nature and timing of the appointment |
| --- | --- | --- | --- | --- |
| Being invited via email or text |  |  |  | .775 |
| Being offered a longer appointment |  |  |  | .689 |
| Having a conversation with the doctor or nurse before the appointment |  |  |  | .476 |
| Being offered medication that prevents muscle cramps and spasms (antispasmodic) or other medication before the appointment |  |  | .561 |  |
| Being offered a test at a different venue e.g. hospital or sexual health venue |  |  | .495 |  |
| A home visit from a doctor or nurse | .500 |  |  |  |
| Being provided with an ambulance or other transport to the appointment | .669 |  |  |  |
| A doctor or nurse who understands my physical disability |  | .814 |  |  |
| A doctor or nurse who discusses my specific needs |  | .852 |  |  |
| A cervical screening passport (e.g. a description of needs and what has worked/not worked before) |  | .581 |  |  |
| Wheelchair access to the surgery | .796 |  |  |  |
| Assistance with undressing/dressing | .843 |  |  |  |
| A hoist to get me on the bed | .837 |  |  |  |
| An examination couch that can be lowered | .606 |  |  |  |
| A sliding board/transfer board/pat slide to get me on the bed | .802 |  |  |  |
| A smaller speculum |  |  | .555 |  |
| Being allowed to insert the speculum myself |  |  | .727 |  |
| A doctor or nurse who is willing to try different solutions |  | .628 | .407 |  |
| Being offered the left lateral position for the screening test |  |  |  |  |
